# Cost-utility analysis of COVID-19 vaccination strategies for endemic SARS-CoV-2 circulation in Canada

**DOI:** 10.1101/2024.12.06.24318620

**Authors:** Rafael N. Miranda, Alison E. Simmons, Michael W.Z. Li, Gebremedhin B. Gebretekle, Min Xi, Marina I. Salvadori, Bryna Warshawsky, Eva Wong, Raphael Ximenes, Melissa K. Andrew, Beate Sander, Davinder Singh, Sarah Wilson, Matthew Tunis, Ashleigh R. Tuite

**Affiliations:** Centre for Immunization Surveillance and Programs, Public Health Agency of Canada, Ottawa, ON, Canada; Dalla Lana School of Public Health, University of Toronto, Toronto, ON, Canada; Public Health Risk Sciences Division, National Microbiology Laboratory, Public Health Agency of Canada, Guelph, ON, Canada; Department of Mathematics and Statistics, McMaster University, Hamilton, ON, Canada; Department of Pediatrics, McGill University, Montreal, QC, Canada; Department of Epidemiology and Biostatistics, Western University, London, ON, Canada; Department of Medicine (Geriatrics), Dalhousie University, Halifax, NS, Canada; Toronto General Hospital Research Institute, University Health Network, Toronto, ON, Canada; Institute of Health Policy, Management and Evaluation, University of Toronto, Toronto, ON, Canada; Public Health Ontario, Toronto, Ontario, Canada; ICES, Toronto, Ontario, Canada; Department of Community Health Sciences, Max Rady College of Medicine, University of Manitoba, MB, Canada

**Author notes:** Both authors contributed equally to this analysis (Rafael N. Miranda, Alison E. Simmons). **Funding** This research was supported, in part, by a Canada Research Chair in Economics of Infectious Diseases held by Beate Sander (CRC-2022-00362).

## Abstract

**Background:** With shifting epidemiology and changes in the vaccine funding landscape, resource use considerations for ongoing COVID-19 vaccination programs are increasingly important. We assessed the cost-effectiveness of COVID-19 vaccination programs, where eligibility is defined by combinations of age and chronic medical conditions, including a strategy similar to current Canadian recommendations, from the health system and societal perspectives.

**Methods:** We used a static, individual-based probabilistic model simulating medically attended COVID-19 in a population of 1 million people followed over a 15-month time period to estimate costs in 2023 Canadian dollars, quality-adjusted life years (QALYs), and incremental cost-effectiveness ratios (ICERs), discounted at 1.5%. COVID-19 epidemiology, vaccine characteristics, and costs were based on the most recently available data.

**Results:** Annual vaccination for adults aged 65 years and older consistently emerged as a cost-effective intervention, with ICERs less than $50,000 per QALY compared to no vaccination for a range of model assumptions. Adding a second dose for adults aged 65 years and older or expanding programs to include vaccination for younger age groups, including those at higher risk of COVID-19 due to chronic medical conditions, generally resulted in ICERs of greater than $50,000 per QALY. Shifting timing of vaccination programs to better align with periods of high COVID-19 case occurrence could result in biannual vaccination for those aged 65 and older being a cost-effective strategy.

**Conclusions:** COVID-19 vaccination programs may be cost-effective when focused on groups at higher risk of disease. Optimal timing of vaccination could improve the cost-effectiveness of various strategies.

## INTRODUCTION

Since the emergence of SARS-CoV-2 in 2019, COVID-19 has caused a high burden of disease worldwide. As of fall 2024, more than 7 million deaths have occurred globally (1) and more than 60,000 deaths have occurred in Canada (2). Although COVID-19 is no longer a public health emergency of international concern (3), it remains a significant cause of morbidity and mortality, particularly among higher risk groups, including older adults and immunocompromised individuals (4–6).

Vaccination against COVID-19 in Canada saved an estimated 500,000 lives and $220 billion between December 2020 and March 2022 (7). With waning immunity following infection and vaccination and the emergence of new SARS-CoV-2 subvariants (8), the administration of additional vaccine doses remains an important tool for optimizing protection (9). Despite the effectiveness of vaccines for preventing severe COVID-19 outcomes, the cost-effectiveness of broad vaccination programs has become less certain, as SARS-CoV-2 is now circulating in populations with prior immune experience derived from infection and/or vaccination. The benefits of COVID-19 vaccination are not uniform across all population groups; for instance, younger adults and children without comorbidities have a lower risk of severe COVID-19 outcomes, and thus may benefit less from vaccination (10).

Prior to 2025, Canadian provinces and territories have been accessing COVID-19 vaccines procured under federal pandemic investments, with vaccine product costs assumed by the Canadian federal government. Beginning in 2025, provinces and territories will transition to usual procurement pathways and will bear the cost of purchasing COVID-19 vaccines for their populations (11).

Given the shifting epidemiology and the changing vaccine funding landscape in Canada, there is an increased focus on efficient use of limited healthcare resources as provinces and territories consider program requirements and budget impact. An evaluation of the cost-effectiveness of different COVID-19 vaccination strategies can help inform vaccination program design. The objective of this study was to estimate the cost-utility of age- and medical-risk based COVID-19 vaccination strategies in Canada.

## METHODS

### Setting

Several COVID-19 vaccines are approved for use in Canada. Although an expert advisory committee of the federal government provides recommendations for the use of vaccines, the primary responsibility for vaccination program decision-making and implementation falls within provincial/territorial responsibility. For fall 2024, Canada’s National Advisory Committee on Immunization (NACI) strongly recommended annual COVID-19 vaccination for people aged 65 years and older and people aged 6 months to 64 years who are at increased of SARS-CoV-2 infection or severe COVID-19 disease (11). For other groups 6 months to 64 years of age who are at lower risk of severe disease, NACI recommended a discretionary annual dose (10). NACI also has a discretionary recommendation for a second vaccine dose at a 6-month interval (with a minimum interval of 3 months) following the first dose for people aged 65 years and older, adult residents of long-term care homes and other congregate living settings for seniors, and people aged 6 months to 64 years who are moderately to severely immunocompromised (12). Of note, only a subset of the population aged less than 65 years strongly recommended to receive an annual dose are recommended to receive two doses per year. This economic evaluation was conducted to inform forthcoming NACI recommendations for the use of COVID-19 vaccines in Canada as provinces and territories transition to vaccine purchasing for their respective populations.

### Model overview

We conducted a model-based cost-utility analysis of COVID-19 vaccination programs in Canada, estimating the cost-effectiveness of vaccination programs based on age- and medical-risk status. The model time period was from July 2024 to September 2025, to represent an annual vaccination program while also allowing for exploration of alternate program timing. We used a discount rate of 1.5% for long-term costs and outcomes, with costs measured in 2023 Canadian dollars and adjusted using the Canadian Consumer Price Index, where necessary (13). Health outcomes were measured as quality-adjusted life years (QALYs). We assessed incremental cost-effectiveness ratios (ICERs) from both the health system (as presented in the main text) and societal perspectives (presented in the supplementary materials) (14). All analyses were conducted using R (15).

### Model structure

Our static individual-based model of COVID-19 cases requiring medical care followed a closed population of 1 million people stratified by age group and medical-risk status (**Supplementary Figure 1**). It was adapted from a previously described model (16). The age group distribution was based on projections for the Canadian population (17). The presence of one or more chronic medical conditions (CMCs) was used to characterize the proportion of the population at higher risk (HR) for experiencing severe outcomes following SARS-CoV-2 infection (18, 19). The remaining proportion of the population without CMCs was characterized as average risk (AR). The model used monthly time steps.

Model parameters for COVID-19 epidemiology, vaccine characteristics, costs, and health utilities are described in more detail below (**Table 1** and **Table 2**). We obtained parameters from published studies and available data, using Canadian sources when possible, and made assumptions if data were not available. When ranges are provided this indicates parameter values were drawn from distributions; beta distributions were used for probabilities and utilities, and gamma distributions were used for costs.

**Table 1.** Population, SARS-CoV-2 epidemiology, and vaccine characteristics input parameters.

| Parameter | Base | Range | Reference |
| --- | --- | --- | --- |
| Population distribution (%) |  |  |  |
| Less than 5 years | 4.7 | -- | Statistics Canada (17) |
| 5-19 years | 13.9 | -- |  |
| 20-49 years | 42.7 | -- |  |
| 50-64 years | 19.1 | -- |  |
| 65-74 years | 11.1 | -- |  |
| 75+ years | 8.6 | -- |  |
| % of population with one or more chronic medical conditions (CMCs) |  |  |  |
| Less than 5 years | 14.4 | -- | CPCSSN (18); Statistics Canada (32); Assumption |
| 5-19 years | 21.0 | -- |  |
| 20-49 years | 24.9 | -- |  |
| 50-64 years | 45.6 | -- |  |
| 65-74 years | 60.1 | -- |  |
| 75+ years | 72.1 | -- |  |
| Monthly % of annual COVID-19 cases: hospitalization data |  |  |  |
| July | 3.7 | -- | Government of Canada (21) |
| August | 3.7 | -- |  |
| September | 12.5 | -- |  |
| October | 15.2 | -- |  |
| November | 16.0 | -- |  |
| December | 19.0 | -- |  |
| January | 10.4 | -- |  |
| February | 6.1 | -- |  |
| March | 4.7 | -- |  |
| April | 3.0 | -- |  |
| May | 3.7 | -- |  |
| June | 1.9 | -- |  |
| Monthly % of annual COVID-19 cases: Respiratory Virus Detection Surveillance System data |  |  |  |
| July | 5.1 | -- | RVDSS (33) |
| August | 8.4 | -- |  |
| September | 12.4 | -- |  |
| October | 13.4 | -- |  |
| November | 13.2 | -- |  |
| December | 12.4 | -- |  |
| January | 8.7 | -- |  |
| February | 5.5 | -- |  |
| March | 4.2 | -- |  |
| April | 3.7 | -- |  |
| May | 5.7 | -- |  |
| June | 7.4 | -- |  |
| Monthly % of annual COVID-19 cases: model projection |  |  |  |
| July | 2.9 | -- | Transmission model (20) |
| August | 5.3 | -- |  |
| September | 8.7 | -- |  |
| October | 14.2 | -- |  |
| November | 19.6 | -- |  |
| December | 20.8 | -- |  |
| January | 10.8 | -- |  |
| February | 5.9 | -- |  |
| March | 4.2 | -- |  |
| April | 2.9 | -- |  |
| May | 2.5 | -- |  |
| June | 2.1 | -- |  |
| Annual probability of medically-attended COVID-19 treated in an outpatient setting |  |  |  |
| 0-4 years | 0.106 | -- | Transmission model (20) |
| 5-19 years | 0.075 | -- |  |
| 20-49 years | 0.075 | -- |  |
| 50-64 years | 0.106 | -- |  |
| 65+ years | 0.120 | -- |  |
| Relative risk of medically-attended outpatient care in people with one or more CMCs compared to people without a CMC |  |  |  |
| All ages | 1.5 | -- | Assumption |
| Annual incidence of COVID-19-attributable hospitalization per 100,000 population |  |  |  |
| 0-4 years | 33.7 | -- | Transmission model (20) |
| 5-19 years | 11.6 | -- |  |
| 20-49 years | 20.6 | -- |  |
| 50-64 years | 64.0 | -- |  |
| 65+ years | 506.9 | -- |  |
| % of patients hospitalized with confirmed COVID-19 with one or more CMCs |  |  |  |
| 0-4 years | 49.3 | -- | COVID-NET (34) |
| 5-19 years | 53.7 | -- |  |
| 20-49 years | 82.2 | -- |  |
| 50-64 years | 97.3 | -- |  |
| 65+ years | 98.9 | -- |  |
| % of patients hospitalized with COVID-19 requiring ICU admission |  |  |  |
| All ages | 13.2 | 12.7-13.6 | CIHI, 2024 (35) |
| COVID-19 mortality per hospitalization (%) |  |  |  |
| 0-4 years | 2.9 | -- | Transmission model (20);<br>Centers for Disease Control<br>and Prevention (36);<br>Assumption |
| 5-19 years | 2.9 | -- |  |
| 20-49 years | 3.3 | -- |  |
| 50-64 years | 9.5 | -- |  |
| 65-74 years | 13.7 | -- |  |
| 75+ years | 17.9 | -- |  |
| All cause mortality rate per year, per 1,000 population |  |  |  |
| All ages | Age-specific<br>rates | -- | Statistics Canada (37) |
| Outpatient COVID-19 duration (days) |  |  |  |
| All ages | 5 | 1-10 | Centers for Disease Control<br>and Prevention (38);<br>Government of Ontario (39) |
| Length of stay in hospital (days) |  |  |  |
| All ages | 24 | 16.2-48.6 | CIHI, 2024 (35) |
| Length of stay in ICU (days) |  |  |  |
| All ages | 9.4 | 2.6-15.9 | CIHI, 2024 (35) |
| % of patients with medically attended COVID-19 prescribed an antiviral |  |  |  |
| 20+ years | 3.1 | 2.5-3.7* | Kuang et al. 2023 (40) |
| Incremental vaccine effectiveness for COVID-19 infection requiring outpatient medical care, by<br>time since vaccination (%) |  |  |  |
| First 2 months | 50 |  | ACIP (28) |
| 3 to 4 months | 32 |  |  |
| 5 to 6 months | 1 |  |  |
| Incremental vaccine effectiveness for COVID-19 infection requiring hospitalization, by time since<br>vaccination (%) |  |  |  |
| First 2 months | 49 |  | ACIP (28) |
| 3 to 4 months | 43 |  |  |
| 5 to 6 months | 14 |  |  |
| Vaccine effectiveness weight for people with one or more CMCs |  |  |  |
| All ages | 0.8 |  | Assumption |
| Vaccination coverage (%) – fall dose |  |  |  |
| 0-4 years | 5.7 |  | Public Health Agency of<br>Canada (41) |
| 5-19 years | 6.3 |  |  |
| 20-49 years | 9.1 |  |  |
| 50-64 years | 23.8 |  |  |
| 65+ years | 52.7 |  |  |
| % of people receiving fall dose who receive a 6-month booster |  |  |  |
| All ages | 20.9 |  | Public Health Agency of<br>Canada (41); assumption |
| Time to receive vaccine (hours), including travel and waiting time |  |  |  |
| All ages | 2 | 1.5-2.5 | Canada Health Infoway (42); Ray et al. 2015 (43) |
| Vaccine wastage rate (%) |  |  |  |
| All ages | 10 | -- | Ministry of Health and Long-Term Care (44); assumption |
| Adverse events following immunization, per 100,000 doses administered |  |  |  |
| Serious adverse event | 1.1 | -- | Public Health Agency of Canada (45) |
| Duration of adverse event following immunization (days) |  |  |  |
| Serious adverse event | 2 | 1-4 | Lee et al. 2009 (46) |
| Probability of developing post-COVID-19 condition (PCC) among outpatient cases |  |  |  |
| 0-4 years | 0.005 | 0.003-0.006 | ACIP (22); Assumption |
| 5-19 years | 0.007 | 0.005-0.009 |  |
| 20+ years | 0.023 | 0.017-0.029 |  |
| Probability of developing PCC among hospitalized cases |  |  |  |
| 0-4 years | 0.008 | 0.003-0.013 | ACIP (22); Assumption |
| 5-19 years | 0.012 | 0.005-0.020 |  |
| 20+ years | 0.040 | 0.017-0.064 |  |
| Duration of PCC (days) |  |  |  |
| All ages | 365 | -- | CCAHS (47) |
| % of PCC cases that miss school or work | 16 | -- |  |
| Duration of school or work missed due to PCC | 23.3 | -- |  |

\*Range defined as $\pm 20\%$ of the base value

**Table 2.** Cost and utility input parameters.

| Parameter | Base | Range | Reference |
| --- | --- | --- | --- |
| Vaccination cost per dose (\$) | | | |
| Vaccine | 43 | 43-107 | Centers for Disease Control and Prevention (48); Mulcahy et al. 2024 (49); assumption |
| Administration | 18 | 13-22 | O'Reilly et al. 2017 (50) |
| Transportation and storage | 0.64 | -- | Sah et al. 2022 (51) |
| Attributable medical costs per person with COVID-19 not requiring hospitalization (\$) | | | |
| All ages | 295 | 207-383 | Sander et al. 2024 (27) |
| Attributable medical costs per person hospitalized with COVID-19 (\$) | | | |
| Hospitalization without ICU admission | 30,147 | 28,760-31,534 | Sander et al. 2024 (27) |
| Hospitalization with ICU admission | 105,677 | 100,467-110,888 |  |
| Direct medical costs for severe adverse event following vaccination (\$) | | | |
| <65 years | 62 | 48-82 | Sander et al. 2010 (52); CIHI (53); Lee et al. 2009 (46) |
| 65+ years | 66 | 51-87 |  |
| Transportation costs (\$) | | | |
| Cost of two-way travel to vaccination or outpatient care (out of pocket) | 14 | 11-16* | Canada Health Infoway (42, 54) |
| Cost of travel to inpatient care | 417 | 210-623 | NACI (54) |
| Co-pay per prescription (\$) | | | |
| 20+ years | 10 | 5-12 | University of Western Ontario (55); The Commonwealth Fund (56); assumption |
| Out-of-pocket medication costs (\$) | | | |
| Over-the-counter medication costs for medically-attended case | 12 | 3-20 | Federici et al. 2018 (57) |
| Prescription medication costs for severe adverse event following vaccination (<65 years) | 4.19 | 3.14-5.24 | Lee et al. 2009 (46) |
| Antiviral costs (20-64 years) | 1,289 | 1,031-1,547* | CADTH, 2024 (58) |
| Caregiver workdays lost |  |  |  |
| % reduction in productivity | 33 | -- | Keita Fakeye et al. 2023 (59) |
| Labour force participation (%) |  |  |  |
| 5-19 years | 16.9 | -- | Statistics Canada (60) |
| 20-49 years | 87.1 | -- |  |
| 50-64 years | 74.6 | -- |  |
| 65-74 years | 18.7 | -- |  |
| 75+ years | 8.0 | -- |  |
| Caregiver | 88.8 | -- |  |
| Average employment income (\$) | | | |
| 5-19 years | 20,600 | -- | Statistics Canada (61) |
| 20-49 years | 60,598 | -- |  |
| 50-64 years | 70,727 | -- |  |
| 65+ years | 50,058 | -- |  |
| Caregiver (age 25-54 years) | 70,452 | -- |  |
| Background health utility |  |  |  |
| 0-4 years | 0.950 | -- | Assumption |
| 5-19 years | 0.917 | -- | Molina et al. 2023 (23) |
| 20-49 years | 0.880 | -- |  |
| 50-64 years | 0.845 | -- | Yan et al. 2023 (24) |
| 65-74 years | 0.867 | -- |  |
| 75+ years | 0.861 | -- |  |
| QALY loss, outpatient care |  |  |  |
| 0-4 years | 0.00575 | 0.00301-0.0085 | Soare 2023 (25) |
| 0-19 years | 0.00561 | 0.00287- 0.00835 |  |
| 20+ years | 0.0045 | 0.0018-0.0074 |  |
| QALY loss, hospitalization without ICU admission |  |  |  |
| 0-19 years | 0.01875 | 0.0052-0.0324 | Soare 2023 (25) |
| 20+ years | 0.0174 | 0.0038-0.031 |  |
| QALY loss, hospitalization with ICU admission |  |  |  |
| 0-19 years | 0.08341 | 0.0592-0.111 | Mercon 2023 (26) |
| 20+ years | 0.0394 | 0.0231-0.0583 |  |
| QALY loss, PCC following outpatient care |  |  |  |
| 0-4 years | 0.1206 | 0.0899-0.1568 | Assumption |
| 5-20 years | 0.1126 | 0.0830-0.1481 | Mercon 2023 (26) |
| 20+ years | 0.0608 | 0.038-0.0912 |  |
| QALY loss, PCC following infection treated in hospital |  |  |  |
| 0-4 years | 0.1412 | 0.1053-0.1836 | Assumption |
| 5-20 years | 0.1319 | 0.09719-0.1734 | Mercon 2023 (26) |
| 20+ years | 0.0712 | 0.0445-0.1068 |  |
| QALY loss, death |  |  |  |
| 0-4 years | 41.38 | -- | Statistics Canada (37, 62) |
| 5-19 years | 38.15 | -- |  |
| 20-49 years | 29.58 | -- |  |
| 50-64 years | 19.07 | -- |  |
| 65-74 years | 13.14 | -- |  |
| 75+ years | 7.35 | -- |  |
| Quality-adjusted life days lost, adverse event following vaccination |  |  |  |
| Serious adverse event | 1.0 | 0.75-1.25 | Sandmann et al. 2021 (63) |

\*Range defined as $\pm 20\%$ of the base value

### COVID-19 epidemiology and disease history

We used a separate age-stratified dynamic compartmental transmission model calibrated to COVID-19 hospital occupancy (from January 2022 to April 2024) and seroprevalence data (from January 2022 to December 2023) that includes vaccination within this fitting window and incorporates immunity levels (20). We then estimated annual cumulative incidence of symptomatic and hospitalized COVID-19 cases in the absence of any COVID-19 vaccination between July 2024 and September 2025. This provided our “no additional vaccination” counterfactual. Full details of the transmission model are provided elsewhere. Annualized incidence estimates for the no vaccination counterfactual scenario were then used to estimate the impact of vaccination for preventing medically-attended COVID-19 cases using the static cost-effectiveness model. This approach allowed for the incorporation of population immunity due to infection and previous vaccination over the modelled time period.

The proportion of annual COVID-19 cases occurring each month was assumed to follow the monthly distribution of hospitalized cases reported from July 2023 to June 2024 (21), and were also assessed using two alternate distributions (**Supplementary Figure 2**). We estimated costs and QALYs associated with medically attended COVID-19 only, which was defined as requiring one of the following levels of care: outpatient (e.g., health care provider or emergency department visit) or inpatient (e.g., hospital admission, with or without intensive care unit (ICU) admission). We assumed that a proportion of people with medically attended COVID-19 developed post-COVID condition (PCC), with a higher risk among people who were hospitalized (22). We also included costs and QALY losses for people with COVID-19 attributable mortality.

### Utilities

We used age-specific utilities based on EQ-5D-5L index scores for the Canadian population to calculate QALY losses associated with COVID-19 mortality (23, 24). QALY losses associated with other modelled health outcomes were derived from published studies (25, 26) and assumptions.

### Costs

We used long-term health care costs attributable to diagnosed COVID-19 derived from a population-based matched cohort study in Ontario, Canada (27). These cost estimates covered a period of approximately one year following initial diagnosis between January and December 2020 for people treated in either outpatient or inpatient settings and included both acute care and post-acute care costs. Post-acute care costs were assumed to include costs associated with PCC and consequently, additional health care costs for PCC were not included in our analysis.

In the absence of Canadian list prices for COVID-19 vaccines, we used a price of $43 per dose for our base case. This estimate is 40% of the US Centers for Disease Control and Prevention (CDC) public list price of $107 per dose and was based on an unpublished Public Health Agency of Canada analysis of historic data that suggests that Canadian negotiated vaccine prices across all vaccine-preventable diseases are typically 30-50% of US public list prices. We explored vaccine prices of 75% ($80) and 100% ($107) of the CDC list price in scenario analyses. We also included costs associated with vaccine administration, vaccine wastage, and adverse events following immunization (AEFIs). Costs for the societal perspective included patient productivity loss due to COVID-attributable disease and death, vaccination and AEFIs, caregiver productivity loss, and out-of-pocket medical costs. Productivity loss was estimated using the human capital and friction cost methods, using age-specific labour force participation rates and average employment income, as described previously (16).

### Vaccination

Vaccination was assumed to occur over a two-month period. In the base-case analysis, vaccination occurred in October and November and, for those receiving it, a second dose was administered 6 months after receipt of the first dose. Timing of dose administration was varied in scenario analyses. Vaccine coverage was based on Canadian estimates of uptake in the spring 2023 and fall/winter 2023-2024 vaccination campaigns.

We assumed different vaccine effectiveness (VE) and waning values for the outcomes of medically attended outpatient and inpatient cases and lower VE for people at higher risk of COVID-19 disease than those at average risk (**Supplementary Figure 3**). VE was assumed to decline over time, with protection falling to 1% against medically-attended outpatient COVID-19 and to less than 15% COVID-19 requiring inpatient care in months 5 and 6 following vaccination. Protection against all outcomes was 0% by month 7 following vaccination. VE estimates were based on US observational data for the 2023-2024 season (28). Vaccination was assumed to reduce risk of PCC by reducing overall SARS-CoV-2 infection risk; we did not include additional VE for preventing PCC in those who did become infected.

### Vaccination strategies

Although COVID-19 vaccines are authorized for the population aged 6 months and older, the youngest modeled age group included the population aged 0 to 4 years, and we did not model births in the population. For this reason, vaccination strategies that included a population aged under 5 years were assumed to apply to the entire age group.

We evaluated a series of increasingly inclusive annual vaccination strategies that added younger age groups, as follows:

- No additional vaccination beyond the vaccinations that had previously been received
- All aged 65 years and older
- All aged 65 years and older and HR aged 50 to 64 years
- All aged 65 years and older and HR aged less than 65 years
- All aged 50 years and older and HR aged less than 50 years

Each of the four vaccination strategies was evaluated including and excluding a second dose (i.e. biannual vaccination) for the population aged 65 years and older (for a total of eight vaccination strategies in addition to the no vaccination strategy). This approach allowed us to examine the key components of the current strategy based on the incremental costs and effects associated with vaccinating the different population groups and the number of doses for those 65 years of age and older. The strategy of biannual vaccination for people aged 65 years and older and annual vaccination higher risk people aged less than 65 years had features most similar to current NACI recommendations. The final strategy (all people 50 years of age and older along with vaccination for people of less than 50 years at higher risk), was more inclusive than the current NACI guidance for those strongly recommended an annual dose.

### Analysis

For each vaccination strategy, we calculated QALYs and costs associated with the modelled health outcomes. We conducted a sequential analysis to compare ICERs. In a sequential analysis, a strategy is removed if others result in more QALYs gained at lower costs (the eliminated strategy is dominated) or if a strategy would never be the optimal intervention regardless of the cost-effectiveness threshold used (i.e., the eliminated strategy is subject to extended dominance) (29). ICER estimates represent the mean of 2,000 model simulations, with each a simulation based on a unique draw from parameter distributions. The number of simulations was selected to ensure adequate sampling from probability distributions. We also calculated outcomes averted compared to no vaccination and number needed to vaccinate to avert an outpatient case, inpatient case, PCC case, or death. Summary results for health outcomes across strategies represent medians and 95% credible intervals (Crl) from the 2,000 model simulations.

### Sensitivity and scenario analyses

We conducted a probabilistic sensitivity analysis for the base-case analysis to estimate the probability that competing vaccination strategies were cost-effective at varying cost-effectiveness thresholds. We performed a sensitivity analysis to explore the sensitivity of the base-case results to assumed vaccine price. For this analysis we identified the optimal strategy at different vaccine prices, with the optimal strategy identified as that with the largest sequential ICER that was below the specified cost-effectiveness threshold. We also conducted a range of scenario analyses focusing on estimates of vaccine wastage, specific vaccine prices, COVID-19 incidence, monthly distribution of COVID-19 cases, and vaccination program timing relative to peak disease activity. Details of the scenarios are provided in **Table 3**.

**Table 3.** Summary of assumptions used in base-case and scenario analyses.

| Scenario | Scenario assumptions* |
| --- | --- |
| Base case | • \$43 per dose of vaccine (40% of CDC list price)<br>• 10% vaccine wastage<br>• VE for HR population is 0.8-fold the VE for AR population<br>• Median estimate of COVID incidence from transmission model<br>• Monthly distribution of cases estimated from 2023-2024 hospitalization data<br>• Annual dose vaccine protection starts in October/November<br>• Second dose protection starts in April/May, for strategies including a second dose for the population aged 65 and older |
| Higher vaccine price (\$80 per dose) | • \$80 per dose of vaccine (75% of CDC list price) |
| Higher vaccine price (\$107 per dose) | • \$107 per dose of vaccine (100% of CDC list price) |
| Higher vaccine wastage (20%) | • 20% vaccine wastage |
| Higher vaccine wastage (30%) | • 30% vaccine wastage |
| Equal VE for HR and AR groups | • VE for HR population is the same as the VE for AR population |
| Higher COVID-19 incidence | • COVID-19 incidence assumed to be 2-times the base-case value |
| Alternate seasonality (flatter curve) | • Monthly distribution of cases estimated from 2023-2024 Respiratory Virus Detection Surveillance System data |
| Alternate seasonality (larger winter wave) | • Monthly distribution of cases estimated from transmission model |
| Earlier program start | • Annual dose vaccine protection starts in August/September |
| Earlier program start and 4-month interval between doses | • Annual dose vaccine protection starts in August/September<br>• Second dose protection starts in December/January, for scenarios including a second dose for the population aged 65 and older |

\*For each scenario except the base case, any changes from the base case are indicated. All relevant base-case scenario assumptions are provided for reference.
Note: AR = average risk (no chronic medical conditions); HR = higher risk (one or more chronic medical conditions).

## RESULTS

### Impact of vaccination strategies

In the absence of additional vaccination in the population, we estimated 8,665 (95% CrI: 8,616 to 8,713) outpatient cases, 133 (95% CrI: 127 to 140) inpatient cases, 178 (95% CrI: 138 to 225) PCC cases, and 16 (95% CrI: 14 to 18) deaths per 100,000 person-years (Supplementary Table 1).

For all vaccination strategies considered, there was a larger proportional impact of vaccination compared to no vaccination for the prevention of COVID-19 hospitalizations and deaths, compared to the prevention of outpatient and PCC cases (**Figure 1, Supplementary Table 1**). The estimated proportion of outcomes averted increased as program eligibility increased. Compared to no additional vaccination for the population, we estimated that 2.3 to 3.5% of outpatient cases, 7.8 to 8.8% of inpatient cases, 2.7 to 4.1% of PCC cases, and 8.7 to 9.6% of deaths could be averted for the vaccination strategies considered based on recent coverage estimates for 2023/2024. Number needed to vaccinate increased for the prevention of each inpatient case, PCC case, or death as program eligibility was expanded to include younger ages. Number needed to vaccinate to prevent an outpatient case remained low even when younger ages were included. Number needed to vaccinate increased when a second dose for adults 65 years and older was included compared to only annual vaccination for this age group.

**Figure 1.**
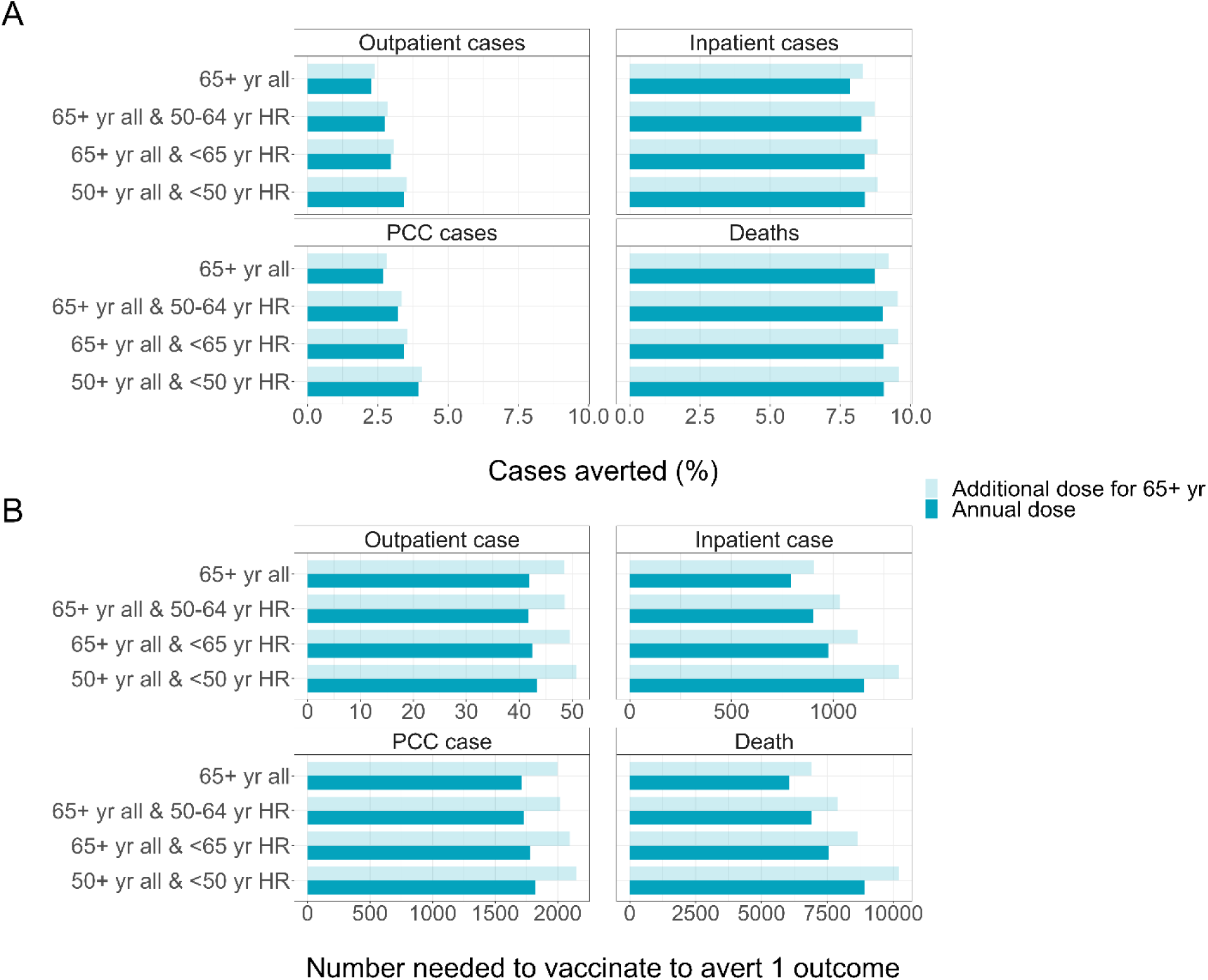
Impact of vaccination on COVID-19 health outcomes. (A) Cases averted for different vaccination strategies compared to no additional vaccination based on COVID-19 coverage data for spring 2023 and fall 2023/2024. (B) Number needed to vaccinate to avert COVID-19 health outcomes for different vaccination strategies. Darker bars denote annual vaccination for the indicated groups while lighter bars denote a second vaccine dose (at a 6-month interval) offered to the population aged 65 years and older and annual vaccination for all other specified groups. Note different x-axis scales for number needed to vaccinate estimates depending on the outcome assessed. Note: HR = higher risk (one or more chronic medical conditions).

### Base-case cost-effectiveness analysis

In the base-case analysis, we found that compared to no additional vaccination beyond those doses received in the past, annual vaccination for all adults aged 65 years and older resulted in an ICER of $7,828 per QALY (**Figure 2**, **Supplementary Tables 2, 3, and 4**). Expansion to include an annual dose for higher-risk adults aged 50 to 64 years resulted in an ICER of $69,399 per QALY compared to an annual dose for adults aged 65 years and older. The next most cost-effective strategy was the addition of a second dose for the 65 years and older age group, with an ICER of $137,505 per QALY compared to annual vaccination for 50 to 64 year olds at higher risk and all those 65 years and older. Finally, expansion to include annual vaccination for the population at higher risk aged less than 50 years resulted in an ICER of $279,975 per QALY compared to a biannual dose for adults aged 65 years and older and an annual dose for higher-risk adults aged 50 to 64 years. Adding the population at average risk aged 50 to 64 years (for whom there is currently only a discretionary, not a strong, recommendation) to receive annual COVID-19 vaccination resulted in an ICER of $529,907 per QALY compared to the current recommendation of biannual vaccination for all 65 years of age and older and annual vaccination for those at higher risk aged less than 65 years.

**Figure 2.**
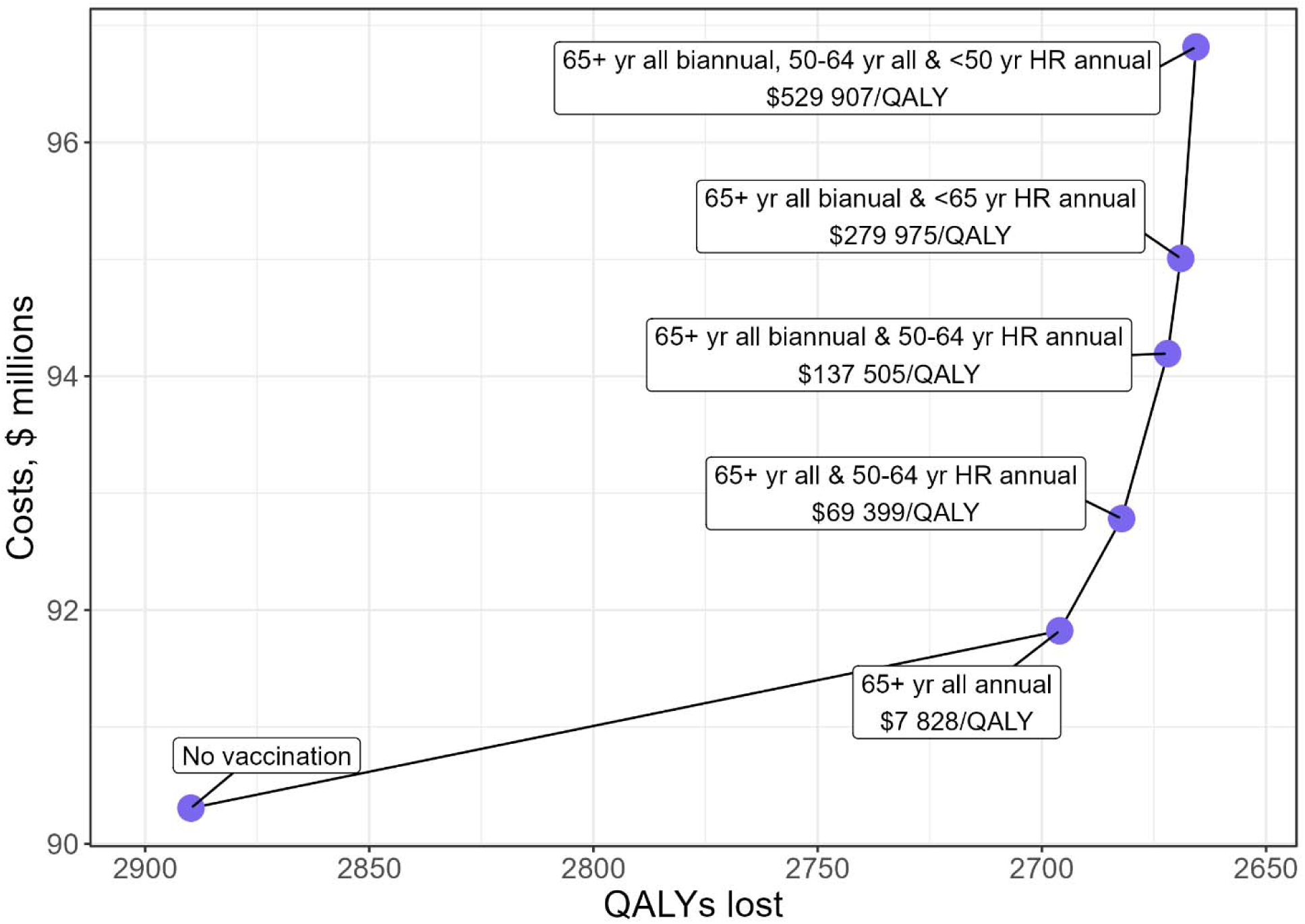
Costs, quality-adjusted life year (QALY) losses, and incremental cost-effectiveness ratios (ICERs) associated with COVID-19 vaccination strategies. Dominated or extended dominated strategies are excluded. The solid line shows the cost-effectiveness frontier that connects non-dominated strategies. Labels show the strategies and sequential ICERs for each strategy, with the ICER calculated relative to the prior (less costly) strategy on the frontier. Results are shown for the base-cases analysis for the health system perspective. Note: HR = higher risk (one or more chronic medical conditions).

Probabilistic sensitivity analysis showed that at a $50,000 per QALY cost-effectiveness threshold, annual vaccination for adults aged 65 years and older had the largest probability of being cost-effective (**Supplementary Figure 4**).

Using a cost-effectiveness threshold of $50,000 per QALY, we found that annual vaccination for all adults aged 65 years and older remained the optimal strategy when vaccine price was between $32 and $110 per dose (**Figure 3**). Vaccine price would need to be between $12 and $31 per dose for annual vaccination of all adults 65 years and older and higher-risk adults aged 50 to 64 years to be the optimal strategy. A vaccine price of less than $12 per dose was required for biannual vaccination for all adults 65 years and older plus annual vaccination for higher-risk adults 50 to 64 years to be the optimal strategy.

**Figure 3.**
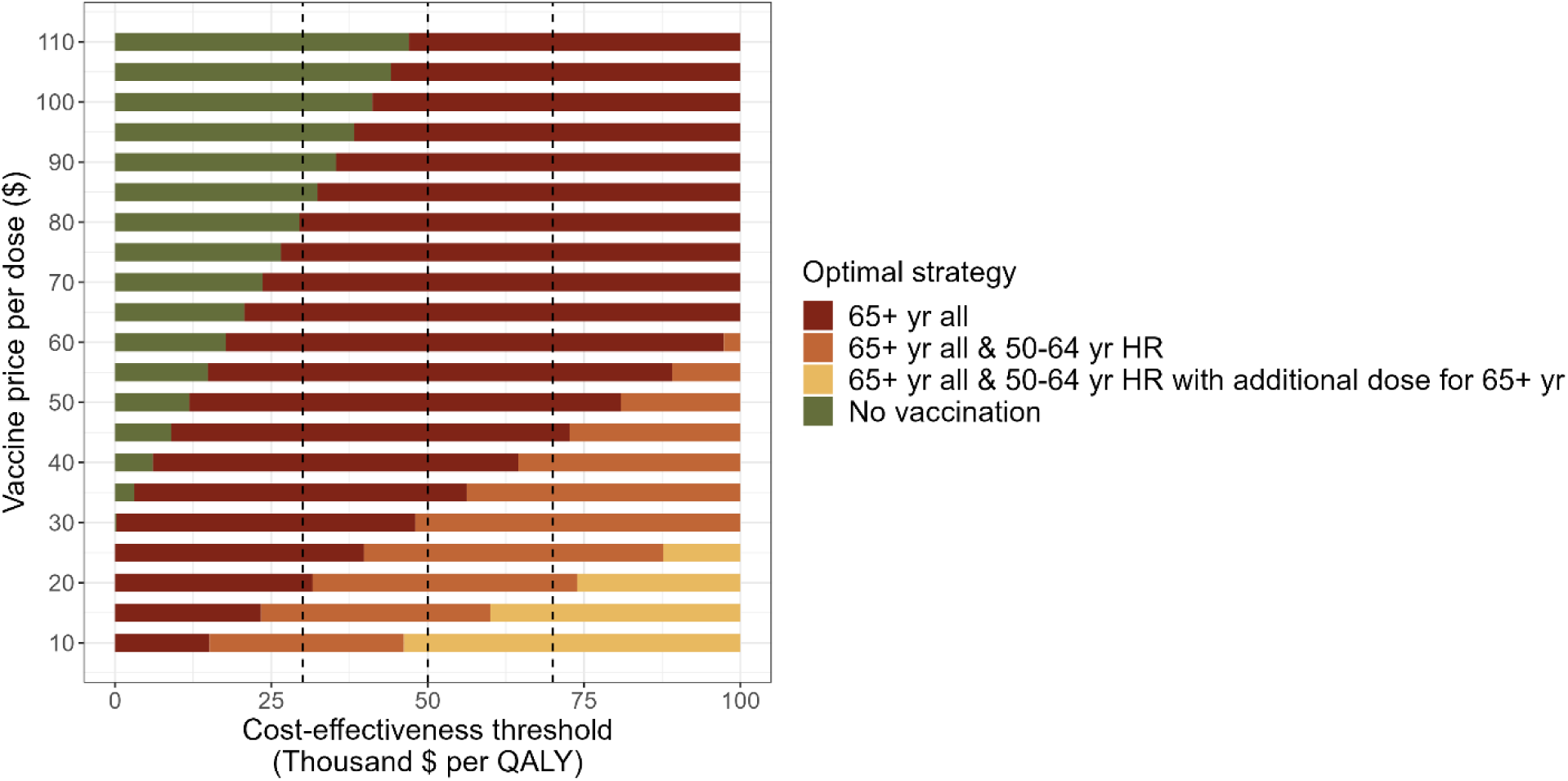
Optimal vaccination strategy for a range of vaccine price and cost-effectiveness thresholds. The optimal strategy is the strategy with a sequential incremental cost-effectiveness ratio less than or equal to the cost-effectiveness threshold and is denoted by the colour of the bar for the indicated vaccine price per dose. The base-case analysis assumed a vaccine price of $43 per dose. For reference, dashed vertical lines indicated cost-effectiveness thresholds of $30,000, $50,000, and $70,000 per QALY. Note: HR = higher risk (one or more chronic medical conditions).

### Scenario analyses

The ICER for annual vaccination for adults aged 65 years and older compared to no additional vaccination remained less than $50,000 per QALY across all scenarios considered, including if vaccine price was increased from $43 per dose included in the base case to $107 per dose (**Table 4**).

**Table 4.** Sequential incremental cost-effectiveness ratios (ICERs) for all vaccination strategies across various scenario analyses.

| Strategy | Change from previous strategy | Sequential ICER (\$ per QALY) | | | | | | | | | | |
| --- | --- | --- | --- | --- | --- | --- | --- | --- | --- | --- | --- | --- |
| | | Base case | Higher vaccine price (\$80 per dose) | Higher vaccine price (\$107 per dose) | Higher vaccine wastage (20%) | Higher vaccine wastage (30%) | Equal VE for HR and AR groups | Higher COVID-19 incidence | Alternate seasonality (flatter curve) | Alternate seasonality (larger winter wave) | Earlier program start | Earlier program start and 4-month interval between doses |
| No vaccination | -- | -- | -- | -- | -- | -- | -- | -- | -- | -- | -- | -- |
| 65+ yr all annual (A) | Annual dose for 65+ yr | 7,828 | 29,471 | 45,264 | 10,115 | 12,401 | 1,154 | 7,979 | 17,460 | 4,923 | 7,116 | 7,024 |
| 65+ yr all & 50-64 yr HR annual (B) | Add annual dose for 50-64 yr HR (to A) | 69,399 | 130,250 | 174,656 | 75,828 | 82,257 | 49,821 | 67,469 | D | 59,860 | 69,918 | D |
| 65 yr all biannual (C) | Add 6-mo dose for all 65+ yr (to A) | D | D | D | D | D | D | D | 44,278 | D | D | 49,418 |
| 65+ yr all biannual & 50-64 yr HR annual (D) | Add 6-mo dose for 65 yr all (to B) or 50-64 yr HR annual (to C) | 137,505 | 239,892 | 314,607 | 148,322 | 159,139 | 103,998 | 120,218 | 118,299 | 188,006 | 134,136 | 88,910 |
| 65+ yr all<br>biannual &<br><65 yr HR<br>annual (E) | Add <50<br>yr HR<br>annual<br>(to D) | 279,975 | 481,131 | 627,921 | 301,227 | 322,480 | 208,259 | 271,466 | 352,241 | 250,083 | 282,375 | 256,887 |
| 65+ yr all<br>biannual,<br>50-64 yr all<br>& <50 yr<br>HR annual | Add 50-64 yr AR<br>(to E) | 529,907 | 885,301 | 1,144,643 | 567,455 | 605,002 | 552,549 | 548,001 | 718,679 | 505,301 | 589,384 | 576,314 |
| 65+ yr all<br>& <65 yr<br>HR annual | -- | ED | ED | ED | ED | ED | ED | ED | D | ED | ED | D |
| 50+ yr all<br>& <50 yr<br>HR annual | -- | D | D | D | D | D | D | D | D | ED | D | D |

Note: AR = average risk (no chronic medical conditions); HR = higher risk (one or more chronic medical conditions); D = dominated; ED = extended dominated.

By contrast to the results for the strategy of annual vaccination for adults aged 65 years and older, and consistent with the base-case analysis, all other vaccination strategies resulted in sequential ICERs greater than $50,000 per QALY in scenario analyses, with three exceptions. First, if COVID-19 incidence had alternate seasonality with a flatter curve, such that infection was less concentrated in the winter months and a higher proportion of cases occurred in the spring and summer months than assumed in the base-case analysis, offering a second vaccine dose for adults aged 65 years and older resulted in an ICER of $44,278 per QALY compared to annual vaccination for this age group. Second, for a scenario assuming an earlier program start and shorter (4-month) interval between doses for those receiving two doses, such that vaccine protection is highest for the 8 contiguous months of highest disease activity, adding a second dose for the population aged 65 years and older resulted in an ICER of $49,418 per QALY compared to annual vaccination for this age group. Finally, if VE is equivalent in high risk and average risk populations, the ICER for annual vaccination of all adults aged 65 years and older and higher-risk adults aged 50 to 64 years was $49,821 compared to annual vaccination for all adults aged 65 years and older.

## DISCUSSION

Our economic evaluation of COVID-19 vaccination programs in Canada found that annual vaccination for the population aged 65 years and older was a cost-effective intervention, even in scenario analyses using more pessimistic assumptions. The next most efficient strategy tended to be program expansion to include adults aged 50 to 64 years with one or more chronic medical conditions. However, in most scenarios, ICERs for including this group exceeded $50,000 per QALY, a commonly used cost-effectiveness threshold.

Our results were sensitive to assumptions about program timing relative to the annual distribution of COVID-19 cases. In scenarios where we assumed less concentration of cases in the winter months or altered program timing to better align with periods of high COVID-19 activity, a program of two doses per year for the population aged 65 and older was identified as the optimal strategy, using a $50,000 per QALY threshold. This likely reflects the high burden of severe COVID-19 in the 65 year and older age group, the waning of vaccine protection over approximately a 6-month period, and the associated benefits of a second dose for preventing disease when vaccination protection is better aligned with periods of high COVID-19 disease incidence.

Our results are consistent with economic evaluations conducted by other National Immunization Technical Advisory Groups that also found that COVID-19 vaccination programs for population members at high risk for severe disease were the most cost-effective. Economic evaluations to support fall 2025 and spring 2026 recommendations by the Joint Committee on Vaccination and Immunisation in the United Kingdom supported universal vaccination for all adults over a specific age cut-off, all residents of care homes for older adults, and all immunosuppressed individuals aged 6 months and older (30). At a combined vaccine price and delivery cost of $44 CAD, all adults 75 years and older would be included in the age-based strategy at a cost-effectiveness threshold of $35,226 CAD (£20,000) per QALY. The final age cut-off for the universal older adult strategy will be determined by vaccine price, which is currently unknown. An economic analysis presented to the American Committee on Immunization Practices (ACIP) estimated ICERs for zero, one, or two doses a year of updated COVID-19 vaccine from the societal perspective (31). For adults aged 65 years and older, the ICER for one dose was $80,443 CAD per QALY compared to no vaccination and the ICER for two doses was $487,311 CAD per QALY compared to one dose. Among individuals aged 5 years to 64 years, ICERs ranged from $284,068 CAD per QALY to $457,892 CAD per QALY for a one dose strategy compared to no vaccination and over $1,000,000 CAD per QALY for a two dose strategy compared to a one dose strategy. The base case adult vaccine price in the model presented to ACIP was $182 CAD per dose, compared to $43 in our model. At similar vaccine prices, ICERs were lower in the model presented to ACIP compared to our model for the strategy of vaccinating adults aged 65 years and older with one annual dose (i.e. a one dose strategy dominated no vaccination from the societal perspective in the model presented to ACIP when the price per dose was $41 CAD). The incidence of COVID-19 attributable hospitalization in the model presented to ACIP was higher than in ours; a higher burden of disease makes vaccination more economically favourable.

Although we conducted extensive scenario analyses to address uncertainty associated with model assumptions, our analysis had some limitations. Use of a transmission model for the no vaccination counterfactual allowed us to estimate expected COVID-19 population burden in a manner that accounted for the impact of continued exposure to SARS-CoV-2 and associated immunity and waning. However, by modelling vaccine protection using a static model, any indirect benefits of vaccination are excluded. Given the low assumed vaccine coverage (based on data from previous years), we expect most of the benefit of vaccination to be due to direct protection for those receiving the vaccine. Our analysis focussed on medically attended cases, which underestimates the societal impacts of infections that do not require medical care but result in work and school absenteeism and associated caregiver impacts. Similarly, by focusing on paid productivity, we are not capturing older adults’ productivity in the informal labour market; as a result, societal perspective cost-effectiveness values may be conservative estimates of the true benefit for vaccination for people aged 65 and older. The assumed decline in VE over 7 months, informed by real world evidence, represents a combination estimate of waning immunity and variant evolution resulting in lower effectiveness against new circulating strains. Given the unpredictability of viral evolution, it is possible that VE could wane faster in the situation of a rapid and significant variant mutation. In the absence of Canadian vaccine price data, we used US CDC public list prices for mRNA vaccines and historical data on relative price differentials between CDC public list prices and Canadian negotiated vaccine prices. We used vaccine price for mRNA vaccines since they represented the majority of vaccines currently used in Canada. It should be noted that applying this historical discount to the CDC public list price for the Nuvaxovid protein subunit vaccine would result in a vaccine price of $34 per dose, which was close to the vaccine price at which program expansion to include 50 to 64 years olds at higher risk of COVID-19 resulted in ICERs less than $50,000 per QALY in our analysis. Finally, the population groups included in our model focused only on key features of the current Canadian recommendations and do not include all those currently recommended for vaccination. In NACI recommendations, beyond people with chronic medical conditions, there are several other groups 6 months to less than 65 years of age, including individuals in or from First Nations, Inuit and Métis communities, members of racialized and other equity-denied communities, pregnant women and pregnant people, and people who provide essential community services who are strongly recommended to receive annual vaccination but were not included in this analysis (11). As well, adult residents of long-term care homes and other congregate living settings for seniors and people aged 6 months to 64 years who are moderately to severely immunocompromised are recommended to receive biannual vaccination based on a NACI discretionary recommendation (12), but these groups were also not included in our analysis.

In summary, our model-based economic evaluation suggests that despite changing epidemiology, COVID-19 vaccination programs in Canada remain cost-effective interventions when focused on populations at higher risk of COVID-19. Alignment of vaccine administration with periods of increased SARS-CoV-2 transmission may increase the efficiency of vaccination programs.

## Data Availability

All data produced in the present work are contained in the manuscript.

## Acknowledgements

The authors are grateful to members of the National Advisory Committee on Immunization COVID-19 Working group for feedback provided during model development and analysis.

