## Supplementary material for "Cost-utility analysis of COVID-19 vaccination strategies for endemic SARS-CoV-2 circulation in Canada"

**Supplementary Figure 1.** Model health states and transitions between states. Boxes indicate health states included in the model and arrows indicate possible transitions between health states. Transitions from any of the health states to death due to background mortality were also included in the model but arrows are not shown. Risk of experiencing medically attended COVID-19 health outcomes were dependent on age, medical risk status, and vaccination status.

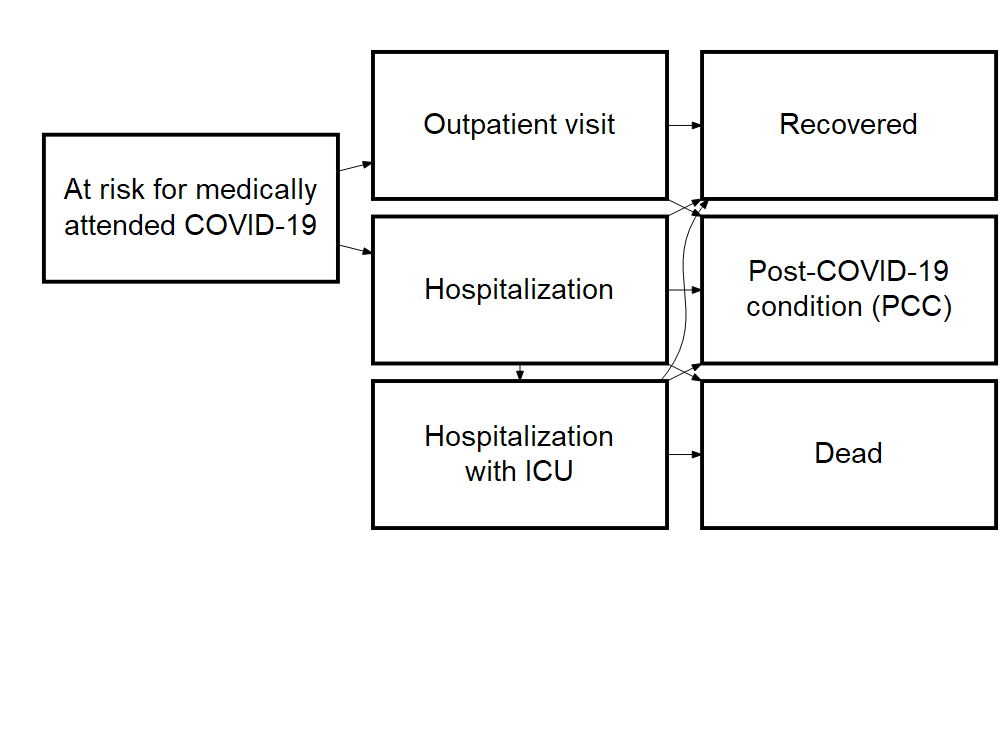

**Supplementary Figure 2.** Assumed annual distributions of COVID-19 cases used in the analysis. The percent of annual COVID-19 cases occurring each month was assumed to follow hospitalization data in the base case analysis. Alternate monthly distributions obtained from the Respiratory Virus Detection Surveillance System (RVDSS) and a transmission model were used in scenario analyses. Hospitalization and RVDSS estimates are based on data for July 2023 to June 2024. The model estimates are based on a projection period covering July 2024 to June 2025.

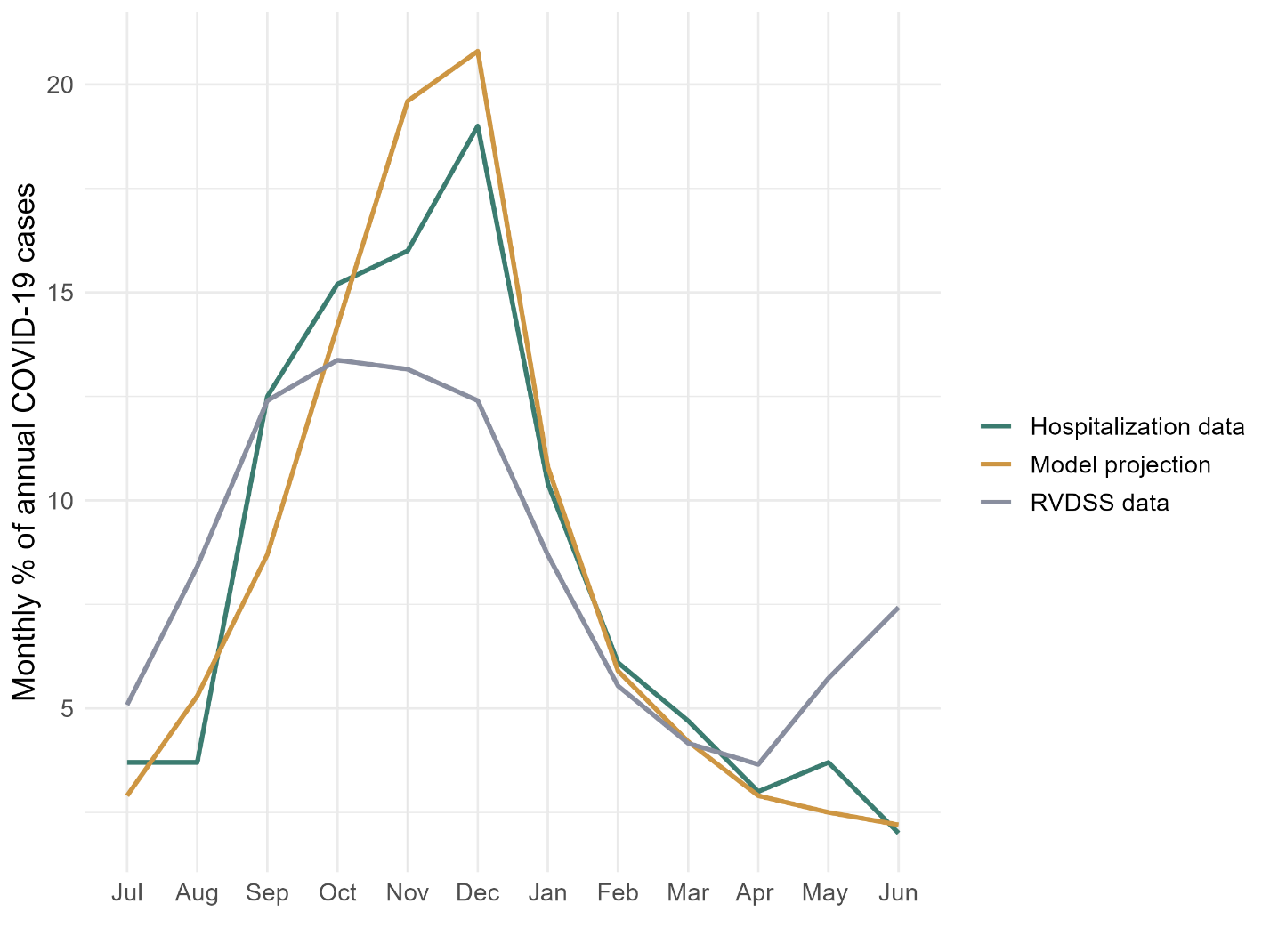

**Supplementary Figure 3.** Incremental vaccine effectiveness (VE) assumptions used in the cost-effectiveness analysis. Different VE was assumed for protection against medically attended COVID-19 requiring outpatient or inpatient care. VE was assumed to be lower for people with chronic medical conditions placing them at higher risk of COVID-19 than for those at average risk in the base-case analysis. Incremental VE was assumed to wane over time, falling to 0 by seven months following vaccination. Note that outpatient VE is assumed to be 1% for months 5 and 6 following vaccination. For people receiving two COVID-19 vaccines a year, receipt of the second dose was assumed to return VE to initial values, with similar waning over time. The graph below assumes a six-month interval between doses, as was assumed in the base case analysis.

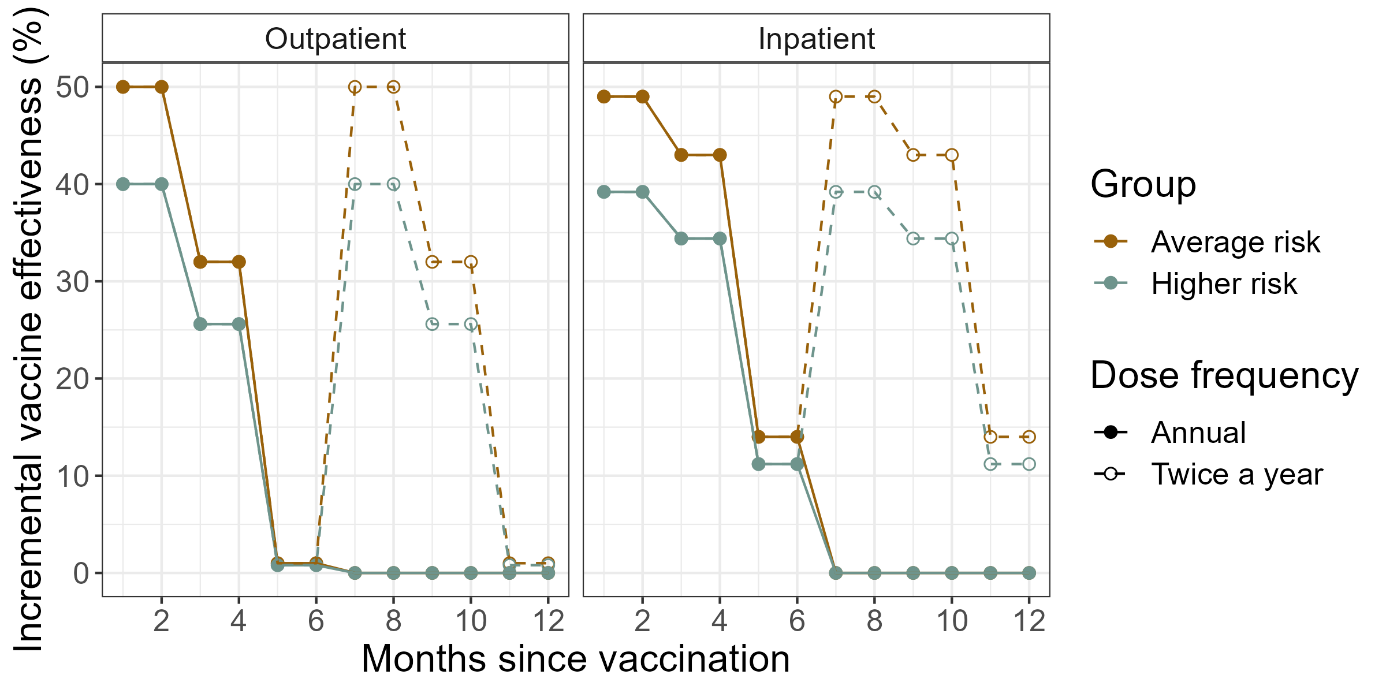

**Supplementary Figure 4.** Cost-effectiveness acceptability curve for the base case probabilistic sensitivity analysis. Points show the proportion of samples for which a given vaccination strategy was identified as cost-effective using a given cost-effectiveness threshold for the (A) health system and (B) societal perspectives based on 2,000 model simulations. The frontier identifies the vaccination strategy with the highest expected net benefit for each threshold value; of note, the optimal strategy based on expected net benefit (as indicated by the frontier) does not always correspond to the strategy with the highest probability of being cost-effective. Results are only shown for vaccination strategies with a probability of cost-effectiveness of 0.1 or greater. Productivity loss for the societal perspective was calculated using the human capital method. Note: HR = higher risk (one or more chronic medical conditions).

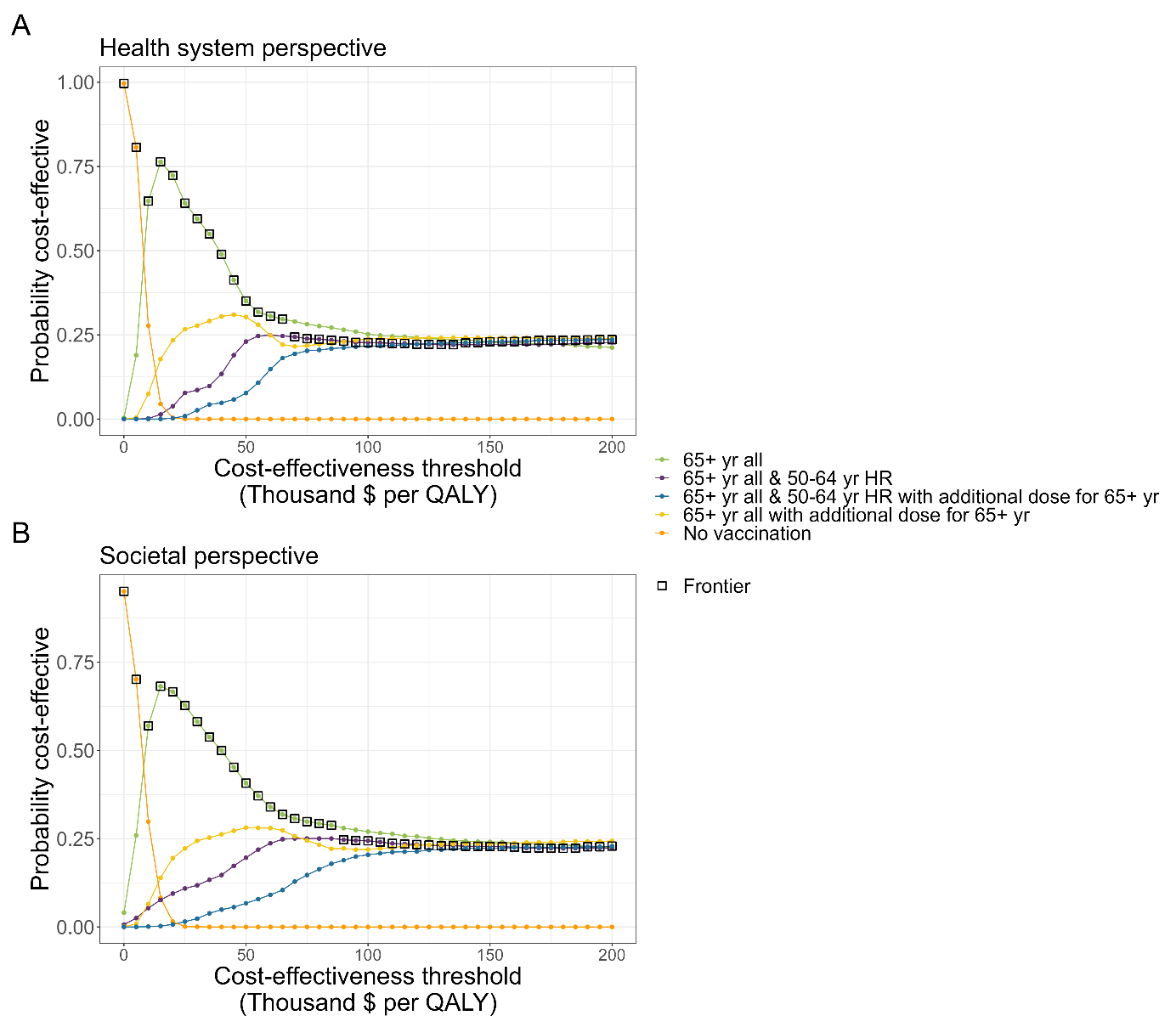

**Supplementary Table 1.** Health outcomes by vaccination strategy, median and 95% credible interval.

| Strategy | Second dose for 65+ yr? | Cases per 100,000 person-years | | | | Cases averted compared to no vaccination (%) | | | | | Number needed to vaccinate to avert one outcome | | | |
| --- | --- | --- | --- | --- | --- | --- | --- | --- | --- | --- | --- | --- | --- | --- |
|  |  | Outpatient | Inpatient | PCC | Death | Outpatient | Inpatient | PCC | Death | Outpatient | | Inpatient | PCC | Death |
| No vaccination | -- | 8665 | 133 | 178 | 16 | -- | -- | -- | -- | -- | | -- | -- | -- |
|  |  | (8616 - 8713) | (127 - 140) | (138 - 225) | (14 - 18) |  |  |  |  |  | |  |  |  |
| 65+ yr all | No | 8467 | 123 | 173 | 14 | 2.3 | 7.8 | 2.7 | 8.7 | 42 | | 792 | 1712 | 6058 |
|  |  | (8418 - 8515) | (117 - 130) | (135 - 219) | (12 - 16) | (2.2 - 2.4) | (6.4 - 9.4) | (2.0 - 3.4) | (5 - 12.8) | (40 - 44) | | (659 - 966) | (1254 - 2510) | (3952 - 11374) |
|  | Yes | 8458 | 122 | 173 | 14 | 2.4 | 8.3 | 2.8 | 9.2 | 48 | | 905 | 1999 | 6905 |
|  |  | (8409 - 8505) | (116 - 129) | (135 - 219) | (12 - 16) | (2.3 - 2.5) | (6.9 - 9.8) | (2.1 - 3.5) | (5.4 - 13.7) | (47 - 50) | | (761 - 1104) | (1447 - 2902) | (4609 - 12438) |
| 65+ yr all & 50-64 yr HR | No | 8427 | 122 | 172 | 14 | 2.7 | 8.3 | 3.2 | 9.0 | | 42 | 902 | 1727 | 6890 |
|  |  | (8378 - 8473) | (116 - 129) | (134 - 218) | (12 - 16) | (2.7 - 2.8) | (6.8 - 9.8) | (2.5 - 4) | (5.3 - 13.2) | (40 - 43) | | (757 - 1102) | (1266 - 2474) | (4582 - 12367) |
|  | Yes | 8418 | 122 | 172 | 14 | 2.9 | 8.7 | 3.4 | 9.5 | 49 | | 1034 | 2021 | 7886 |
|  |  | (8369 - 8465) | (115 - 129) | (134 - 218) | (12 - 16) | (2.8 – 3.0) | (7.2 - 10.2) | (2.6 - 4.1) | (5.5 - 14) | (47 - 50) | | (878 - 1252) | (1464 - 2936) | (5343 - 13658) |
| 65+ yr all & <65 yr HR | No | 8408 | 122 | 172 | 14 | 3.0 | 8.4 | 3.4 | 9.0 | 42 | | 976 | 1782 | 7547 |
|  |  | (8359 - 8455) | (116 - 129) | (134 - 218) | (12 - 16) | (2.9 - 3.1) | (6.9 - 9.9) | (2.7 - 4.2) | (5.4 - 13.2) | (41 - 44) | | (822 - 1187) | (1319 - 2515) | (5020 - 13554) |
|  | Yes | 8399 | 122 | 172 | 14 | 3.1 | 8.8 | 3.6 | 9.6 | 50 | | 1120 | 2098 | 8640 |
|  |  | (8350 - 8446) | (115 - 129) | (134 - 218) | (12 - 16) | (3.0 - 3.2) | (7.4 - 10.3) | (2.8 - 4.4) | (5.5 – 14.0) | (48 - 51) | | (949 - 1357) | (1516 - 2987) | (5858 - 14957) |
| 50+ yr all & <50 yr HR | No | 8367 | 122 | 171 | 14 | 3.4 | 8.4 | 4.0 | 9.0 | 43 | | 1152 | 1822 | 8916 |
|  |  | (8319 - 8414) | (116 - 129) | (133 - 216) | (12 - 16) | (3.3 - 3.5) | (6.9 - 9.9) | (3.1 - 4.8) | (5.4 - 13.2) | (42 - 45) | | (970 - 1404) | (1359 - 2551) | (5932 - 16007) |
|  | Yes | 8359 | 122 | 171 | 14 | 3.5 | 8.8 | 4.1 | 9.6 | 51 | | 1322 | 2151 | 10205 |
|  |  | (8310 - 8405) | (115 - 129) | (133 - 217) | (12 - 16) | (3.4 - 3.7) | (7.4 - 10.3) | (3.3 - 4.9) | (5.5 - 14) | (49 - 52) | | (1121 - 1605) | (1577 - 3039) | (6918 - 17683) |

Note: HR = higher risk (one or more chronic medical conditions); QALY = quality-adjusted life year; ICER = incremental cost-effectiveness ratio.

**Supplementary Table 2.** Costs, QALYs, and sequential ICERs for base case scenario by strategy: health system perspective.

| **Strategy** | **Costs ($)** | **Effect (QALYs lost)** | **Incremental Costs ($)** | **Incremental Effect (QALYs gained)** | **Sequential ICER** |
| --- | --- | --- | --- | --- | --- |
| No vaccination | 90,305,263 | 2,889.792 | -- | -- | -- |
| 65+ yr all annual | 91,822,017 | 2,696.031 | 1,516,754 | 193.7606 | 7,828 |
| 65+ yr all & 50-64 yr HR annual | 92,781,484 | 2,682.206 | 959,468 | 13.8254 | 69,399 |
| 65+ yr all biannual & 50-64 yr HR annual | 94,192,769 | 2,671.942 | 1,411,284 | 10.2635 | 137,505 |
| 65+ yr all biannual & <65 yr HR annual | 95,007,415 | 2,669.033 | 814,646 | 2.9097 | 279,975 |
| 65+ year all biannual, 50-64 yr all & <50 yr HR annual | 96,816,201 | 2,665.619 | 1,808,786 | 3.4134 | 529,907 |
| 65+ yr all & <65 yr HR annual | 93,432,115 | 2,679.163 | -- | -- | Extended dominated |
| 65+ yr all biannual | 92,945,261 | 2,686.134 | -- | -- | Dominated |
| 50+ yr all & <50 yr HR annual | 94,901,680 | 2,675.781 | -- | -- | Dominated |

Note: HR = higher risk (one or more chronic medical conditions); QALY = quality-adjusted life year; ICER = incremental cost-effectiveness ratio.

**Supplementary Table 3.** Costs, QALYs, and sequential incremental ICERs for base case scenario by strategy: societal perspective (human capital method).

| **Strategy** | **Costs ($)** | **Effect (QALYs lost)** | **Incremental Costs ($)** | **Incremental Effect (QALYs gained)** | **Sequential ICER** |
| --- | --- | --- | --- | --- | --- |
| No vaccination | 196,626,076 | 2,889.792 | -- | -- | -- |
| 65+ yr all annual | 198,068,598 | 2,696.031 | 1,442,522 | 193.7606 | 7,445 |
| 65+ yr all & 50-64 yr HR annual | 199,270,659 | 2,682.206 | 1,202,062 | 13.8254 | 86,946 |
| 65+ yr all biannual & 50-64 yr HR annual | 201,179,391 | 2,671.942 | 1,908,731 | 10.2635 | 185,972 |
| 65+ yr all biannual & <65 yr HR annual | 202,351,295 | 2,669.033 | 1,171,904 | 2.9097 | 402,756 |
| 65+ year all biannual, 50-64 yr all & <50 yr HR annual | 205,156,498 | 2,665.619 | 2,805,204 | 3.4134 | 821,820 |
| 65+ yr all & <65 yr HR annual | 200,161,866 | 2,679.163 | -- | -- | Extended dominated |
| 65+ yr all biannual | 199,471,633 | 2,686.134 | -- | -- | Dominated |
| 50+ yr all & <50 yr HR annual | 202,358,450 | 2,675.781 | -- | -- | Dominated |

Note: HR = higher risk (one or more chronic medical conditions); QALY = quality-adjusted life year; ICER = incremental cost-effectiveness ratio.

**Supplementary Table 4.** Costs, QALYs, and sequential ICERs for base case scenario by strategy: societal perspective (friction cost method).

| **Strategy** | **Costs ($)** | **Effect (QALYs lost)** | **Incremental Costs ($)** | **Incremental Effect (QALYs gained)** | **Sequential ICER** |
| --- | --- | --- | --- | --- | --- |
| No vaccination | 177,972,315 | 2,889.792 | -- | -- | -- |
| 65+ yr all annual | 180,085,082 | 2,696.031 | 2,112,767 | 193.7606 | 10,904 |
| 65+ yr all & 50-64 yr HR annual | 181,528,768 | 2,682.206 | 1,443,686 | 13.8254 | 104,423 |
| 65+ yr all biannual & 50-64 yr HR annual | 183,478,935 | 2,671.942 | 1,950,167 | 10.2635 | 190,009 |
| 65+ yr all biannual & <65 yr HR annual | 184,715,092 | 2,669.033 | 1,236,157 | 2.9097 | 424,839 |
| 65+ yr all biannual & 50-64 yr all & <50 yr HR annual | 187,530,329 | 2,665.619 | 2,815,237 | 3.4134 | 824,759 |
| 65+ yr all biannual | 181,521,235 | 2,686.134 | -- | -- | Extended dominated |
| 65+ yr all & <65 yr HR annual | 182,489,702 | 2,679.163 | -- | -- | Extended dominated |
| 50+ yr all & <50 yr HR annual | 184,695,465 | 2,675.781 | -- | -- | Dominated |

Note: HR = higher risk (one or more chronic medical conditions); QALY = quality-adjusted life year; ICER = incremental cost-effectiveness ratio.
